# Mortality and predictors of Chronic Kidney Diseases (CKD) in selected dialysis centers in Addis Ababa, Ethiopia

**DOI:** 10.1101/2022.01.27.22269879

**Authors:** Beza Zewdu Desta, Behailu Tariku Derseh, Abel Fekadu Dadi

**Author notes:** Corresponding author: Beza Zewdu Desta. Email: Behailu Tariku Derseh Abel Fekadu Dadi.

## Abstract

**Background:** Chronic Kidney Disease (CKD) affects 10-15% of the population worldwide. The number of dialysis patients is steadily rising in Ethiopia. Chronic hemodialysis patients experience high rates of mortality; however, data is lacking in Ethiopia. We explored the mortality and its driving factors in selected dialysis center in Addis Ababa

**Method:** we retrospectively followed a group of End-Stage Renal Disease patients on hemodialysis from 2016 to2020 in St. Paul Millennium Medical College (SPMMC), Zewditu Memorial Hospital (ZMH), and Menellik II Hospital. We fitted Kaplan Meier analysis to estimate the one and five year’s survival probability of these patients and Cox Proportional regression analysis to model the predictors of mortality at p value ≤0.05.

**Result:** We analyzed a data of (139) patients under follow up. The mean age (± SD) of the patients was 36.8 (±11.95) years. Twenty-four (17%) of the patients died at the end of follow-up. The median survival time was 46.2 months (95% CI: 41.8, 50.5). The one and five-year’s survival probability of these patients was 64.8% and 18.7%, respectively. Our analysis showed that patients with hypertension (AHR = 4.33; 95% CI: 1.02, 34.56), cardiac complication (AHR = 4.69; 95% CI: 1.32, 16.8), and infection during dialysis (AHR = 3.89; 95% CI: 1.96, 13.8) were more likely to die.

**Conclusion:** The survival status of hemodialysis patients in three was low. Preventing and treating comorbidities and complication during dialysis would reduce mortality of CKD patients.

## Introduction

Chronic kidney disease (CKD) is a major public health problem that challenging health system around the world(1). Chronic Kidney disease (CKD) is a progressive loss of kidney function over a period of months or years. Professional guidelines classified the severity of CKD in five stages. A clinically significant Stage 5 CKD or end-stage renal disease is the most deliberating stage in which the patient should get Renal Replacement Therapy(RRT) (2).

A death associated with CKD has nearly be doubled worldwide between 1990 and 2010 and was the 18 highest cause of death in 2010 (3). It is estimated that by 2030 more than 70% of patients with end-stage renal disease will be living in low-income countries (4).

Renal replacement therapy (RRT) is the mainstay of care for patients with an end stage renal disease(ESRD) to reduce mortality and morbidity (5).

Patients with end-stage renal diseases (ESRD) are unable to sustain life without dialysis support(6). That would last about 3 to 4 hours a week. The duration of each session depends on how well the patient’s kidneys work, and how much fluids weight the patient has gained between treatments (6).There are three main types of vascular accesses (VA) used in HD treatment: Arterio-Venous Fistula (AVF), Synthetic Arterio Venous Graft (AVG), and Central Venous Catheter (CVC) (7). All types of VA have their own risks and expensive. The clinical practice guidelines recommend, AVF as first choice because of its reduced associated complications, morbidity, and mortality compared with the AVG and CVC(8).

Although there is limitation of data about prevalence of CKD, few studies indicate that renal disease has become a significant public health problem in Ethiopia. A cross sectional study showed that the prevalence of CKD in Ethiopia is estimated to be 12.2% and increased in the last few years following an increasing prevalence of diabetes and hypertension. The prevalence of CKD is as high as 41.0% in age <35 years and 62% in males (9).

The high rate of mortality among dialysis patients after initiation of the therapy is attributed to factors such as demographic, comorbidities, blood markers such as albumin and hemoglobin, and type of vascular access (10)

Duration of dialysis per session, hypertension, and infection status significantly affect the survival rate of ESRD. (11).

A study on survival pattern of hemodialysis patient in Ethiopia shows that 45.1 % of death occurs during dialysis treatment and 23.1 % of the patient died within the first 90 days of initiation of dialysis, while only 42.1% of the patients survive more than a year. Septicemia (34.1%), cardiovascular disease (29.3%) and uses of the catheter as vascular access was associated with a short and long term survival of patients treated with hemodialysis (5).

There is limited data reporting mortality and its predictors among chronic hemodialysis kidney patients that we intended to investigate in this study.

## Methodology

### Study setting and period

We analyzed a retrospectively recorded data of patients enrolled in a maintenance hemodialysis in three hospitals in Addis Ababa (St. Paul Millennium Medical College (SPMMC), Zewditu Memorial Hospital (ZMH), and Menelik II) from 1^*st*^January 2016 to 30^*th*^ December 2020.

Addis Ababa has a population size of 3,435,028 of whom 1,809,577 are females (12).There are 13 publics and 34 privates hospitals in the centers providing different public health services.

We included all chronic kidney patients age 18 years and above who were on hemodialysis for End-Stage Renal Disease during the study period.

We excluded: (1) Patients who starts hemodialysis for acute renal failure or patients who started dialysis on an emergency basis or acute dialysis (with the duration from first to last dialysis being less than 30 days); (2) those with incomplete medical record/charts for important variables; and (3) alive patients transferred into COVID-19 center with their medical cards as these patients had restricted access.

### Outcome of interest

The outcome is death of patients happening during the time of hemodialysis and we both interested in modeling the mortality and time. Right censored patients were those alive, default, and transferred during the time of follow up. The start of the follow-up was the time the patient started maintenance hemodialysis. We calculated an incidence rate by considering the time of follow-up of each patient contribute to the exposure time.

### Covariates

We did a literature review to identify the potential risk factors associated with death. The covariates included: (1) Sociodemographic such as age, sex, marital status, residence; (2) weight in kg, comorbidities such as hypertension, diabetes mellitus, cancer, chronic kidney disease-mineral and bone disorder (CKD-MBD), types of vascular access for dialysis (AV Fistula, Grift, Catheter); (3) cardiovascular disease such as coronary artery disease (CAD), cardiac arrhythmia, cardiac failure, cardiac valvular disease, pericardial disease, cardiomyopathy, and congenital heart disease; (4) biochemical profile of the patients such as Serum levels of hemoglobin, creatinine, albumin, Phosphorus; (5) duration of dialysis per season and frequency of dialysis per week.

### Data source

We extracted the required data from medical records and hemodialysis registration books using a pretested data extraction form. The investigator further checked for the validation and completeness of the extracted data. The variables included in this study were recorded in an Excel file. Missing values for important variables were completed by reviewing clinical notes and reports available in the selected dialysis center. Trained two Masters of Public Health Students and two Bachelor of nursing from each hospital, and one supervisor were involved in the data extraction and validation. Both supervisor and data collectors were trained for two days on how to extract data, what to be extracted and to make them internalize the context of each question in the data extraction form.

### Data analysis

We entered the extracted data into Epi data and exported to SPSS for further analysis. We used mean or median and standard deviation to present numeric data and frequencies and percentages to present a categorical data.

We computed the person-time of follow-up from the date of starting hemodialysis to death, loss to follow-up, or the end of the study. We calculated the Incidence rate of death by dividing the number of deaths among CKD patients occurring by person-months of follow-up. We used actuarial life table to estimate survival after initiation of hemodialysis; and the Kaplan-Meier test to estimate the probability of death and the median time to death after initiation of hemodialysis. We did log-rank test to compare a time to death between different covariates. We checked assumption of proportional hazard model using the Log (-log (St)) plots and tests. We used the Cox proportional hazard model to determine the probability of death after initiation of hemodialysis adjusting for confounding factors. We excluded covariates that violate the assumption of cox proportional hazard model from the analysis. All statistically significant variables having a p-value ≤ 0.25 in the bivariate analysis were adjusted in the final model. We estimated the crude and adjusted hazard ratio (HR) and its 95% confidence interval (CI) were estimated to identify and report significant predictors of death at p-value < 0.05.

## Results

We found total of 139 chronic dialysis patients who started a dialysis between January 1, 2016, and December 30, 2020.

### Socio demographic characteristic of the patients

Table 1 presents the characteristics of patients included in this study. Slightly above half of the patients were males 88 (63.3%). The mean (±SD) age at dialysis initiation was 36.81(±11.50) years and more than half were in younger age category (18-34 years) (show table 1).

**Table 1:** Socio Demographic characteristics of 139 ESRD patients on maintenance hemodialysis for in Addis Ababa Ethiopia, from January 1st 2016 to December 31st 2020.

| Characteristic |  | Frequency(n) | Percentage |
| --- | --- | --- | --- |
| Age (mean ±SD<br>(35.81±11.9)) | 18-34 | 77 | 55.4 |
|  | 35-54 | 48 | 34.5 |
| | ≥55 | 14 | 10.1 |
| Gender | Male | 88 | 63.3 |
|  | Female | 51 | 36.7 |
| Occupation | Government | 25 | 18.0 |
|  | Private | 30 | 21.6 |
|  | Unemployed | 57 | 41.0 |
|  | Farmer | 1 | 0.7 |
|  | Student | 6 | 4.3 |
|  | Not recorded | 20 | 14.4 |
| Residence | Addis Ababa | 102 | 73.4 |
|  | Out of Addis<br>Ababa | 37 | 26.6 |
| Marital status | Single | 59 | 42.4 |
|  | Married | 75 | 54.0 |
|  | Divorced | 3 | 2.2 |
|  | Widow | 2 | 1.4 |

Majority of the patients 102(73.4%) were living in Addis Ababa while the rest 26.6 % (37) came to Addis for treatment from different regions of Ethiopia(show table 1).

### Baseline characteristics of the patients

The mean (±SD) of systolic blood pressure was 143.25 mmHg (±22.063) at the initiation of dialysis. The mean of blood pressure at the last session of dialysis was 142.10 (±19.083 mmHg). The mean (±SD) weight of the patients at dialysis initiation of dialysis was 55.54 kg (show table 2).

**Table 2:** Base line data blood pressure and weight with their statistical parameter ESRD patients on maintenance hemodialysis for in Addis Ababa Ethiopia, from January 1st 2016 to December 31st 2020.

| Base line data of the patient | Mean | Median | Standard division | Range | Minimum | Maximum |
| --- | --- | --- | --- | --- | --- | --- |
| Systolic BP(first) | 143.95 | 140 | 21.935 | 129 | 92 | 221 |
| Diastolic BP (first) | 87 | 90 | 14.941 | 73 | 53 | 126 |
| Systolic BP(last) | 142.1 | 140 | 19 | 99 | 95 | 194 |
| Diastolic BP(last) | 84 | 86 | 13 | 86 | 44 | 130 |
| Weight (first) | 55.54 | 54 | 11.42 | 56.1 | 31.8 | 87.9 |
| Weight(last) | 54.9 | 53 | 10.99 | 57 | 30 | 87 |

**Table 3:** Patient Laboratory value with their statistical parameter ESRD patients on maintenance hemodialysis for in Addis Ababa Ethiopia, from January 1st 2016 to December 31st 2020.

| Serum electrolyte (first and last) | Mean | Median | Standard deviation | Range | Minimum | Maximum |
| --- | --- | --- | --- | --- | --- | --- |
| Creatinine(mg/dl) | 8.9 | 8.6 | 3.55 | 18.0 | 2 | 20.04 |
| Hemoglobin | 10.37 | 10.1 | 2.27 | 11.1 | 5 | 16.1 |
| Hematocrit | 31.81 | 6.98 | 43 | 32 | 15.8 | 47.8 |
| Phosphorus | 2.21 | 1.7 | 1.59 | 7.47 | 0.43 | 7.9 |
| Potassium | 4.83 | 4.92 | 1.0 | 6.97 | 0.85 | 7.82 |
| Calcium | 3.39 | 2.23 | 2.4 | 8.7 | 0.92 | 9.61 |
| Albumin | 3.86 | 3.9 | 0.8 | 4.8 | 2.1 | 6.9 |
| Sodium | 136.7 | 138 | 6.59 | 48 | 99 | 147 |
| Urea | 104.2 | 96.8 | 46.1 | 202.6 | 10.4 | 213 |
| Creatinine(mg/dl) | 5.14 | 4.32 | 3.03 | 14.0 | 0.6 | 14.6 |
| Hemoglobin | 10.2 | 10.2 | 1.87 | 10 | 5 | 15 |
| Hematocrit | 31.35 | 31 | 6.12 | 32.5 | 15 | 47.5 |
| Phosphorus | 2.0 | 1.7 | 1.3 | 7.9 | 0.52 | 8.3 |
| Potassium | 4.95 | 4.9 | 0.99 | 4.8 | 2.9 | 7.7 |
| Calcium | 3.65 | 2.4 | 2.6 | 9.2 | 1.2 | 10.4 |
| Albumin | 3.5 | 3.5 | 0.54 | 3.0 | 2 | 5 |
| Sodium | 137.3 | 137.0 | 4.14 | 22 | 127 | 149 |
| Urea | 57 | 46 | 39 | 158.75 | 5.25 | 16.4 |

### Treatment modality

Most of the patients (89.2%) had three sessions of dialysis per week. Duration of each session ranged between three to four hours. Most of the patients 121(87.1%) had four hours of stay in dialysis.

More than half (63.3%) of the hemodialysis patients were taking their treatment by Arteriovenous Fistula (AVF) followed by catheter (AVC) 36%.

About more than 80% of the patients were given some form of medication for complications of CKD and hemodialysis treatment and 82.2% of the patient were taking Erythropoietin (EPO). Twenty -four (17.3 %) of the patients had been given a blood transfusion at least once during their time on dialysis. The most common causes of ESRD were hypertension (48.2%), diabetes mellitus (6.5%), Glomerulonephritis (6.5%), Renal Stone (2.2 %) and 36.7% of the ESRD patients had unknown etiology (show table 4 and 5).

**Table 4:** Treatment modalities for 139 ESRD patients on maintenance hemodialysis for in Addis Ababa Ethiopia, from January 1st 2016 to December 31st 2020.

| Variables | Characteristic | N (%) | Percentage |
| --- | --- | --- | --- |
| Frequency of dialysis session/week | Twice | 15 | 10.8 |
|  | Three | 124 | 89.2 |
| Duration of dialysis per session | Three hours | 5 | 3.6 |
|  | Three and half hours | 13 | 9.4 |
|  | Four hours | 121 | 87 |
| Types of vascular access | Catheter | 50 | 36 |
|  | Arteriovenous fistula | 88 | 63.3 |
|  | Arteriovenous graft | 1 | 0.7 |
| Cause of CKD | Hypertension | 67 | 48.2 |
|  | Diabetes mellitus | 9 | 9 |
|  | Glomerulonephritis | 9 | 9 |
|  | Renal stone | 3 | 3 |
|  | Unknown | 51 | 51 |
| Blood transfusion | Yes | 24 | 17.3 |
|  | No | 115 | 82.7 |
| EPO treatment | Yes | 124 | 89.2 |
|  | No | 15 | 10.8 |
| Cause of death | Cardiovascular disease | 6 | 25 |
|  | Infection | 4/24 | 16.7 |
|  | Uremic complication | 3/24 | 12.3 |
|  | Sudden death | 4/24 | 16.7 |
|  | Hyperkalemia | 1/24 | 4.1 |
|  | Cardiac arrest | 2/24 | 8.3 |
|  | Cardiovascular disease | 4/24 | 16.7 |

**Table 5:** patient status for 139 ESRD patients on maintenance hemodialysis for in Addis Ababa Ethiopia, from January 1st 2016 to December 31st 2020.

| Variable | Category | Total (%) | Number of event (%) | Censored (%) |
| --- | --- | --- | --- | --- |
| Sex | Male | 88(63.3) | 15(17) | 73(83) |
|  | Female | 51(36.7) | 9(17.6) | 42(82.4) |
| Resident | Addis Ababa | 102(73.4) | 19(18.63) | 83(81.37) |
|  | Out of Addis | 37(26.6) | 5(13.51) | 32(86.49) |
| Marital status | Married | 75(54.7) | 12(16) | 63(84) |
|  | Unmarried | 63(43.3) | 12((19) | 51(81) |
| Vascular access | AVF | 88(63.3) | 6(6.8) | 82(93.2) |
|  | Catheter | 50(36) | 17(34) | 33(66) |
|  | Graft | 1(0.7) | 1(100) | 0(0) |
| Frequency of dialysis per week | Twice per week | 15(10.8) | 7(46.7) | 8(53.3) |
|  | Three per week | 124(89.2) | 17(13.7) | 107(86.3) |
| Duration of dialysis | Three hours | 5(3.6) | 2(40) | 3(60) |
|  | Three and half hour | 13(9.4) | 4(30.8) | 9(69.2) |
|  | Four hours | 121(87) | 18((14.9) | 10385.1) |
| Hypertension | Yes | 96(69) | 22(23) | 74(77) |
|  | No | 43(31) | 2(4.65) | 41(95.35) |
| Diabetes mellitus | Yes | 11(7.9) | 3(27.3) | 8(72.7) |
|  | No | 128(92.1) | 21(16.4) | 107(83.6) |
| albumin(mg/dl) | Under 3.5 | 57(41) | 18(31.6) | 39(68.4) |
|  | 3.51-4.00 | 45(32.4) | 2(4.4) | 43(95.6) |
|  | Above 4.01 | 37(26.6) | 4(10.8) | 33(89.2) |

**Table 6:** unadjusted survival HR (95% CI) of ESRD patients on maintenance hemodialysis for in Addis Ababa Ethiopia, from January 1st 2016 to December 31st 2020.

| Variable | Category | p-value | Crude HR (95%CI) |
| --- | --- | --- | --- |
| Hypertension | Yes | 0.047 | 4.331(1.018-18.431) * |
|  | No* |  | 1 |
| Types of vascular access | Graft | 0.036 | 10.069(1.166-86.976) |
|  | Catheter | .000 | 9.479(3.558-25.24) *** |
|  | Fistula* |  | 1 |
| Infection | Yes | 0.038 | 3.625(1.071-12.255) * |
|  | No* |  | 1 |
| Cardiovascular complication | Yes | 0.015 | 4.495(1.338-15.105) * |
|  | No* |  | 1 |
| Frequency of dialysis per week | Two times | 0.28 | 2.7(1.116-6.533) |
|  | Three times* |  | 1 |
| The reference categories were those indicated in asterisk (*) |  |  |  |

\*Significant (P-value < 0.05), \*\*significant (p-value<0.01) and\*\*\* significant (p<0.001)
CHR: - Crude Hazard Ratio, AHR: - Adjusted Hazard Ratio, HR=1 is reference variable.

### Comorbidity and complication

Some forms of comorbidities were occurred in 78(56.1%) of the patients. Cardiovascular complication was the most common comorbid complication recorded in 53.2% of the patients as moderate-severe mitral regurgitation and diastolic dysfunction. Seventy-eight (56.1%) of the patients were developed different types of infections sometimes in their course of treatment. For example, Hypotension occurred in (36.0%) of the patients at least once during the period of dialysis (show table 4).

### Survival status of patients on hemodialysis

About 24 (17%) patients died and the rest 115(83%) were censored during the follow up. The sample included 88 male patients of which 15(17%) were died and 51 females of which nine were (17.65%) died. Twenty four patients were died during the first 60 months follow-up. Among these, 10 and 4 were dead during the first and second six months of follow-up, respectively. The median survival time was 46.2 months and the overall survival rate was86% at one year and 70% at five years.

### Predictors of mortality

Hypertension, cardiovascular complication, infection, and types of vascular access were turned-out to predict the mortality in adjusted model (show figure 1 and table 7). As such being hypertensive increased the risk of mortality by 4.3 times (95%CI: 1.02-18.43).

**Table 7:** adjusted survival HR (95% CI) of ESRD patients on maintenance hemodialysis for in Addis Ababa Ethiopia, from January 1st 2016 to December 31st 2020.

| Variable | Category | p-value | Unadjusted HR (95%CI) |
| --- | --- | --- | --- |
| Hypertension | Yes | 0.015 | 7.139(1.474-34.565) |
|  | No* |  | 1 |
| Types of vascular access | Graft | 0.010 | 19.568(2.008-190.736) |
|  | Catheter | 0.09 | 3.839(1.404-10.498) |
|  | Fistula* |  | 1 |
| Infection | Yes | 0.035 | 3.586(1.985-13.054) |
|  | No* |  | 1 |
| Cardiovascular complication | Yes | 0.017 | 4.696(1.316-16.763) |
|  | No* |  | 1 |
| The reference categories were those indicated in asterisk (*) |  |  |  |

NB: \*Significant (P-value < 0.05), \*\*significant (p-value<0.01) and\*\*\* significant (p<0.001) CHR: - Crude Hazard Ratio, AHR: - Adjusted Hazard Ratio, HR=1 is reference variable.

**Figure 1:**
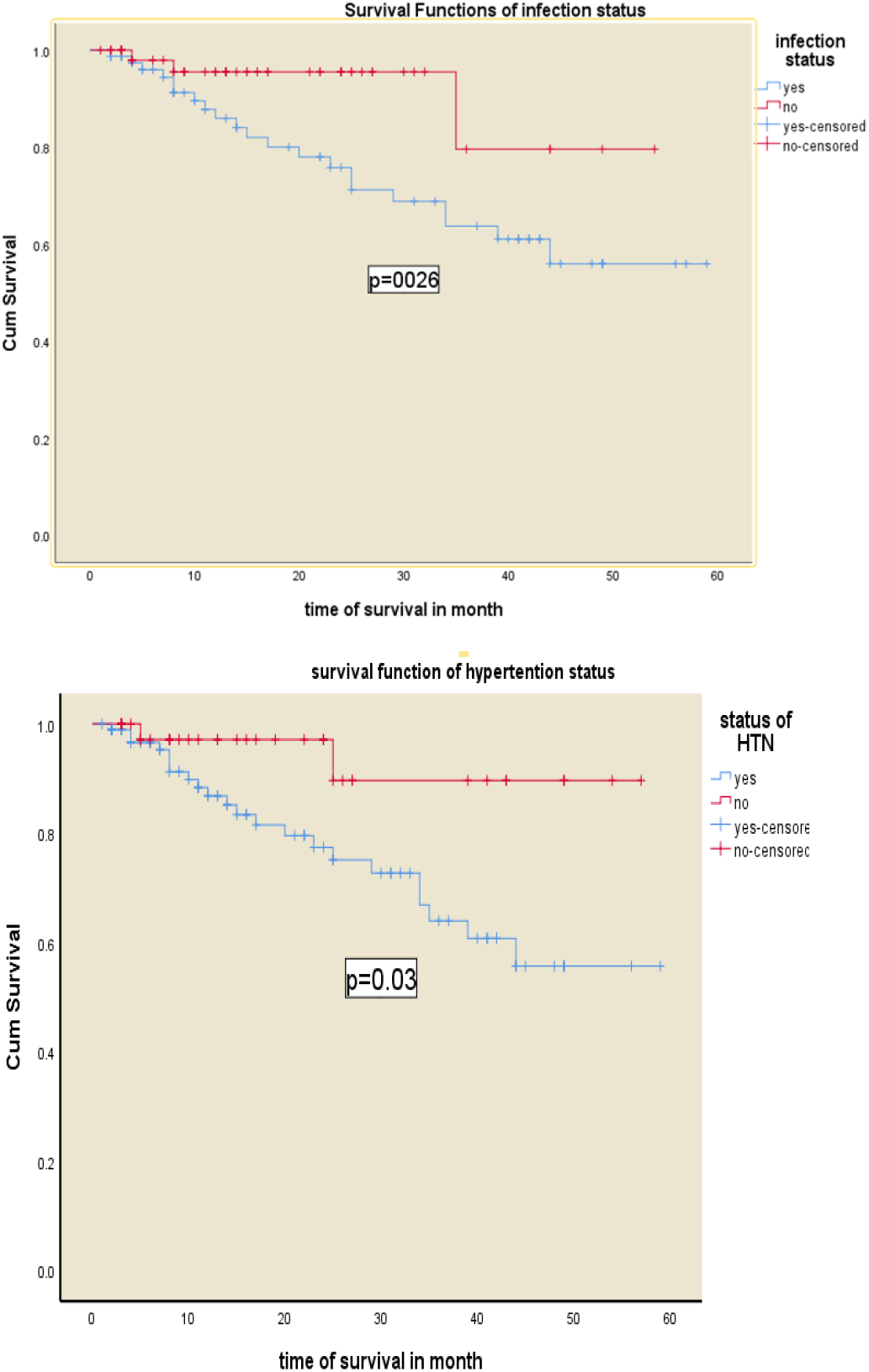
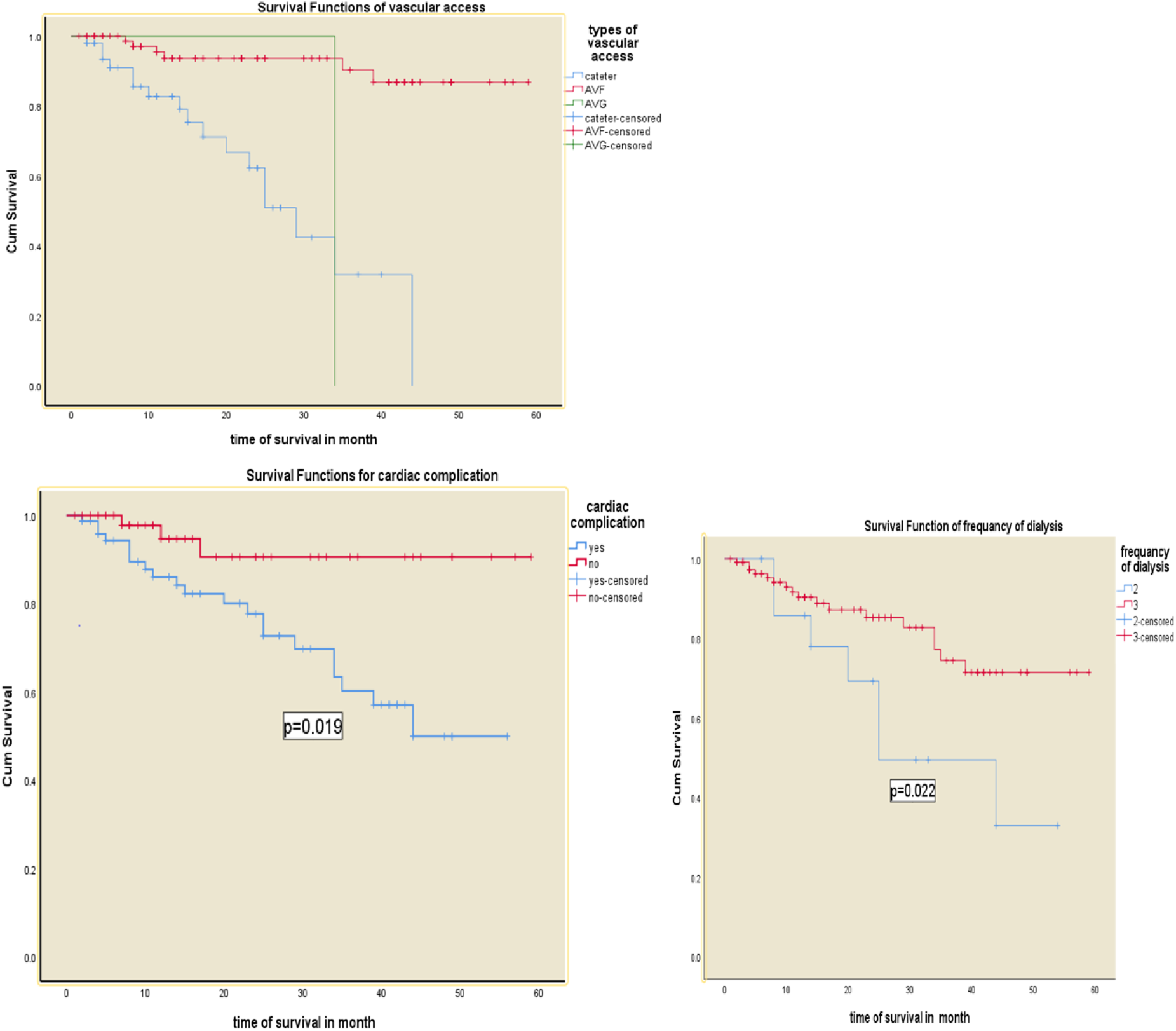
Kaplan-Meier survival curves for hemodialysis patients, according to the mean of the covariate of 139 patients diagnosed in 2016-2020 in, Addis Ababa Ethiopia.

Patients with cardiovascular complication had a 4.7 (95%CI: 1.32-16.76) times higher risk of mortality. The risk of mortality among patients using Arterio-Venous Graft and temporary venous catheter were 19.57 (95%CI: 2.01-190.74) and 3.84(95%CI: 1.40, 10.50) times higher than those patients using Arterio-Venous Fistula. Furthermore, a risk of mortality was 3.59 (95%CI: 1.98-13.05) times higher among patients with comorbid infection.

## Discussion

Dialysis remains the priority intervention for patients with advanced CKD as there are limited organs for transplantation and excesses risks associated with the transplantation for most of patients worldwide. Dialysis prolong and improve quality of life of patients with kidney failure, and this needs a continues evaluation of the procedure to improve treatment outcomes. We aimed to conduct this secondary data analysis as an evaluation and to inform clinicians about factors that trigger the mortality of patients in dialysis centers in Addis Ababa.

The median follow-up time of the patients was 46.2 months and its consistent with patients in Adama hospital that estimated to be 43 months(13). Furthermore, the survival rate of patients under follow up was 70% at 5-years and 86% at 1-years which relatively the same with a similar study reported elsewhere which reported to be 80%(14). Another study from Brazil reported a survival rate of 84.7% in the first and 63.3% in the five years of treatment (15). The main reason for the discrepancy might be the low accessibility and cost of the service in Ethiopia. The lower long-term survival rate of the patients could be attributed to in that undergoing dialysis is very low and can be attributable to medical conditions that we have identified including; types of vascular access type, hypertension, infection status, and cardiac complications.

Patients with cardiovascular complications had around four times increased risk of mortality compared to those patients with no cardiac complications. In another way, the mean survival months of patients with cardiac complication was 14 months lower. This result is similar to a study reported from China; were cardiac complication was a statistically significant predictor of mortality among ESRD patients(16). Similarly, we found a cumulative hazard of mortality that was three times among patient with CVD complications it has been reported that stroke was a prominent predictor of all-causes of mortality in patients who had cardiovascular complications (16). The burden of chronic kidney disease on mortality is significant and continues to increase, which requires, optimal medical management. Studies reported the excess of cardiovascular morbidity and mortality in patients with renal insufficiency particularly (17).

The risk of death for ESRD patients with infection was about three times higher than that than of the patients on hemodialysis without infection. Similar study in Ethiopia found a risk of death in patients in maintenances hemodialysis with septicemia which accounts for 34.1% of all-cause of mortality (5). Another study from Taiwan found a risk which is lower but in similar direction. Having infection and advanced CKD increased the risk from all cause of mortality by 34% in the first year and 19% during the nine years follow up period (18). Efforts at minimizing infectious complications, preventing infection at any level should increase survival among hemodialysis patients. In the present study, another factor that was identified as a predictor of mortality among patients on hemodialysis was the type of vascular access for hemodialysis treatment. Those who were on arteriovenous Catheter had a risk of mortality about four times compared to those patients with AFV access. This result is consistent with a study reported in Iran a 3.6-fold higher risk of death (19). This study showed that a vascular access using catheter accompanied with increased risk of death and this is highly likely as the fact that. In fact, limiting the use of catheters minimized the infection complications in patients.

Another two studies show that catheter use was associated with a mortality risk of 67% and three times as compared with use of an AVF(17). We similarly found a risk of mortality of about 47% higher among patients who used Catheter.

Dissimilar to our result a retrospective cohort study in Belgium found a vascular access type was not independently correlated with patient survival, even after taking into account change of vascular access over time(20). And also the result of this study is consistent with nighty one hemodialysis case summary of hemodialysis patents in Ethiopia revel that patients who had catheter as permanent vascular access was associated with poor outcome as compared to artery venous fistula and Graft and one-year survival rate for AVC group were 5.4 % and 66.9 % in other types of vascular access(p-value <0.0001)(5). In summary most of studies found that Arteriovenous catheter increases the risk of death compared to other vascular access. This might be due to a high risk of infection in patient taking treatment by AVC.

Hypertension were another significant predictor of mortality in risk of having that was four times. High blood pressure in our analysis was associated with a mortality risk of four times and this is inconsistent with studies that found reverse association i.e., high blood pressure associated with low risk and low blood pressure associated with high risk of mortality in dialysis patients. Contradict to this study some of the existing studies refer to this phenomenon as “reverse epidemiology”, indicating a paradoxical association between mortality and the effect of hypertension in dialysis patients. This paradoxical phenomenon of lower BP or a decline in BP over time is associated with increased mortality and higher BP is associated with lower mortality is described as “reverse” epidemiology of hypertension (21).

There are several possible explanations for this finding in the literature, for instance, discrete (binary) categorization of hypertension at the baseline as (yes or no) instead of using blood pressure as a continuous variable (SBP, DBP). Binary categorization of hypertension indeed did not reveal how severe it was at the baseline. This categorization, due to insufficient information on blood pressure measurements, and duration of antihypertensive medication use or any relevant treatment at the baseline could have resulted in reporting normotensive or well-controlled hypertension patients as hypertensive. Similar to our study, an Indonesian finding found a high blood pressure that associated with a high risk of mortality though the risk is low compared to our study (15).

Overall, this study can provide preliminary information about predictors that affect survival status of CKD patients on hemodialysis in the three selected hemodialysis centers in Addis Ababa, Ethiopia. Since we have limited information about this topic our study would have paramount importance in recognizing the problems and complication in hemodialysis patients. However, due to limited sample size of the study we might not confidently rely on this information and we recommend further investigation by overcoming the apparent limitation of this study. Being a retrospective study we were unable to find some important predictors that should be considered in the study leaving a question of potential residual confounding bias.

## Conclusions

The survival status of hemodialysis patients in the three hospitals was low. Preventing and treating comorbidities and complications during dialysis would reduce the mortality of CKD patients.

## Data Availability

All relevant data are within the manuscript and its Supporting Information files

## Computing interest

The authors declare that not have conflict of interest

## Ethics approval and consent to participate

Ethical clearance was obtained from the Ethical Review Committee of Debre Berhan University, College of Health Sciences (Protocol number: 12/21/CHS/SPH). Permission was obtained from all Hospitals and dialysis Centers. Confidentiality and privacy of the information were assured and maintained by preventing disclosure of the information to third parties.

## Acknowledgements

The authors give extended gratitude to Debre Berhan University College of Health Sciences, Department of Public Health for providing the golden opportunity to carry out this study, all staff members of SPHMMC, ZMH, MRH, Data collectors, family and friends who supported in all efforts to accomplish the study.

## References

1. Jha V, Garcia-garcia G, Iseki K, Li Z, Naicker S, Plattner B, et al. Global Kidney Disease 3 Chronic kidney disease : global dimension and perspectives. Lancet [Internet]. 2013;382(9888):260–72. Available from: http://dx.doi.org/10.1016/S0140-6736(13)60687-X

2. Couser W, Remuzzi G, Mendis S, Tonelli M. The contribution of chronic kidney disease to the global burden of major noncommunicable diseases. Kidney Int. 2011 Dec 1;80:1258–70.

3. Lozano R, Naghavi M, Foreman K, Lim S, Shibuya K, Aboyans V, et al. Global and regional mortality from 235 causes of death for 20 age groups in 1990 and 2010: A systematic analysis for the Global Burden of Disease Study 2010. Lancet. 2012;380(9859):2095–128.

4. Sumaili EK, Krzesinski JM, Zinga C V., Cohen EP, Delanaye P, Munyanga SM, et al. Prevalence of chronic kidney disease in Kinshasa: Results of a pilot study from the Democratic Republic of Congo. Nephrol Dial Transplant. 2009;24(1):117–22.

5. Shibiru T, Gudina EK, Habte B, Deribew A, Agonafer T. Survival patterns of patients on maintenance hemodialysis for end stage renal disease in Ethiopia : summary of 91 cases [Internet]. Vol. 14, BMC Nephrology. BMC Nephrology; 2013. 1 p. Available from: BMC Nephrology

6. Lakshmi KR, Nagesh Y, Veerakrishna M. Performance Comparison of Three Data Mining Techniques for Predicting Kidney Dialysis Survivability. Int J Adv Eng Technol. 2014;7(1):242–54.

7. Rezapour M, Zadeh MK, Sepehri MM. Implementation of Predictive Data Mining Techniques for Identifying Risk Factors of Early AVF Failure in Hemodialysis Patients. Hindawi Publ Corp. 2013;2013.

8. Rezapour M, Taran S, Parast MB. The impact of vascular diameter ratio on hemodialysis maturation time : Evidence from data mining approaches and thermodynamics. MJIRI. 2016;

9. Kore C, Tadesse A, Teshome B, Daniel K, Kassa A, Ayalew D. The Magnitude of Chronic Kidney Disease and its Risk Factors at Zewditu Memorial Hospital, Addis Ababa, Ethiopia. J Nephrol Ther. 2018;08(03):8–12.

10. Collins AJ, Foley RN, Chavers B, Gilbertson D, Herzog C, Johansen K, et al. US renal data system 2011 Annual data report. Am J Kidney Dis. 2012;59(1 SUPPL. 1):A7.

11. Mekonen MW, Birahan KA, Chekole DM, Derso EA. Determinants of Overall Survival of Kidney Failure for Patients Receiving Dialysis in Saint Geberial General Hospital, Addis Ababa, Ethiopia. J Kidney OPEN. 2020;1–7.

12. Berhanu G, Tibebu, Jelaludin Ahmed, Abebaw Ferede S, Negera A, Bezu HTA, Kassie K narayana. UG, Abelti G, et al. Population Projections for Ethiopia. In: Central Statistical Agency. 2013.

13. Hussein M, Muleta G, Seyoum D, Kifle D, Bedada D. Survival Analysis of Patients with End Stage Renal Disease the Case of Adama Hospital, Ethiopia Mekiya. Clin Med Res. 2017;6(6):201–8.

14. Lima JJG De, Sesso R, Abensur H, Lopes HF, Giorgi MCP, Krieger EM, et al. Nephrology Dialysis Transplantation Predictors of mortality in long-term haemodialysis patients with a low prevalence of comorbid conditions. Eur Dial Transpl Assoc. 1995;1708–13.

15. Urte Zakauskiene, Alvita Vickiene, Vaidas Vicka, Diana Sukackiene, Laurynas Rimsevicius, Marius Miglinas. Institute of Clinical Medicine, Vilnius University Faculty of Medicine V. E-POSTERS BLOOD PRESSURE DURING HEMODIALYSIS AS PREDICTOR OF. Wolters Kluwer Heal. 2021;2021.

16. Tong J, Liu M, Li H, Luo Z, Huang J, Liu R, et al. Mortality and Associated Risk Factors in Dialysis Patients with Cardiovascular Disease. kidney internatonal. 2016;479–87.

17. Astor BC, Eustace JA, Powe NR, Klag MJ, Fink NE. Type of Vascular Access and Survival among Incident Hemodialysis Patients : The Choices for Healthy Outcomes in Caring for ESRD (CHOICE) Study. Am Soc Nephrol. 2005;1449–55.

18. Chang C, Fan P, Kuo G, Lin Y, Tsai T. Infection in Advanced Chronic Kidney Disease and Subsequent Adverse Outcomes after Dialysis Initiation : A Nationwide Cohort Study. Sci Rep [Internet]. 2020;1–10. Available from: http://dx.doi.org/10.1038/s41598-020-59794-7

19. Salman Khazaei MY, Nematollahi1, ShahrzadZobdeh2, Zahra Sheikh3, VidaMMansournia ohammad A. Survival Rate and Predictors of Mortality among Hemodialysis Patients in West of Iran, 1996 – 2015. Int J Prev Med |. 2018;1–5.

20. Clerck D, Bonkain F, Cools W, Niepen P Van Der. Vascular access type and mortality in haemodialysis : a retrospective cohort study. BMC Nephrol. 2020;1–7.

21. Georgianos PI, Agarwal R. Blood Pressure and Mortality in Long-Term Hemodialysis — Time to Move Forward. Am J Hypertens. 2017;30(March):211–22.

